# Untargeted serum metabolomics reveals novel metabolite associations and disruptions in amino acid and lipid metabolism in Parkinson’s disease

**DOI:** 10.1101/2022.12.29.22284028

**Authors:** Kimberly C Paul, Keren Zhang, Douglas I Walker, Janet Sinsheimer, Yu Yu, Cynthia Kusters, Irish Del Rosario, Aline Duarte Folle, Adrienne M Keener, Jeff Bronstein, Dean P Jones, Beate Ritz

## Abstract

**Objectives:** Recent advances in high-resolution metabolomics platforms allow the simultaneous measurement of thousands of small molecules produced from metabolism (metabolites), providing a map of disease-related perturbations across interconnected pathways. We used high performance, untargeted metabolomics to identify metabolic disturbances and molecular events associated with Parkinson’s disease (PD) in two population-based studies.

**Methods:** We performed a metabolome-wide association study (MWAS) on PD, using serum-based untargeted metabolomics data derived from high resolution liquid chromatography, mass spectrometry (LCMS). We used two independent, case-control populations for discovery and replication (n=642 PD patients, n=277 controls).

**Results:** From the LCMS, 5,145 metabolites were detected across the two study populations in ≥50% of the samples (HILIC: 2913 metabolites; C18: 2063 metabolites). Using logistic regression and an FDR to correct for multiple testing, we determined 236 metabolites were associated with PD in a meta-analysis at an FDR<0.05. Of these, 110 metabolites were independently associated with PD in both discovery and replication studies at p<0.05 (187 at p<0.10), while 24 were associated with levodopa-equivalent dose among the PD patients. Intriguingly, the microbial related metabolite, p-cresol (meta-OR=1.29, 95% CI=1.13, 1.47, FDR=0.01), and it’s two metabolites, p-cresol sulfate and p-cresol glucuronide, were found at higher intensity among the PD patients relative to controls. P-cresol glucuronide was also associated with motor symptoms among patients. Pyroglutamic acid (meta OR=3.79, 95% CI=2.60, 5.54; FDR=5.30E-09), the anti-inflammatory metabolite itaconate (meta OR=0.47, 95% CI=0.36, 0.61; FDR=8.44E-06), and cysteine-S-sulfate (meta OR=1.56, 95% CI=1.32, 1.83; FDR=1.66E-05) were also among the most strongly associated metabolites. Seventeen pathways were also enriched, including several related to amino acid and lipid metabolism.

**Conclusions:** Our results revealed PD-associated metabolites in two independent study populations, implicating individual metabolites including p-cresol and itaconate, as well as suggesting metabolic disturbances in amino acid and lipid metabolism and inflammatory processes.

## Introduction

Parkinson’s disease (PD) is a complex, multi-factorial neurodegenerative disease with multi-system involvement. Pathologically, PD is defined by the loss of dopaminergic neurons in the substantia nigra and widespread intracytoplasmic aggregations of misfolded α-synuclein(1). Rare genetic mutations have been identified in early-onset, familial PD, but idiopathic PD’s complex etiopathogenesis remains unclear(2).

High-throughput technological developments over the past decade have paved the way for agnostic analysis of multiple omic measures, providing insights into disease etiology. Genome-wide association studies (GWAS), for instance, have highlighted the role of endolysosomal (vesicle trafficking, lysosomes, and autophagy) and immune pathways in PD(3). Still, biologic processes are dynamic and operate through complex interactions between gene expression, protein function, and metabolism. Investigating other principal omics, including the metabolome, may provide novel insights into biologic processes involved in PD.

Recent advances in high-resolution metabolomics platforms allow for the simultaneous measurement of thousands of metabolites. Metabolic networks shed light on the underlying biochemical activity of cells, tissues, and organs, enabling a multi-system level approach to the study of PD. Furthermore, metabolites reflect the convergence of genomic, epigenomic, transcriptomic, and proteomic action in tandem with the system’s response to environmental exposures, thus offering a readout of both physiologic and pathologic states of an individual(4). Metabolites circulating in the blood provide a wealth of information about biologic processes across different systems, including the central nervous system as metabolites can cross the blood-brain barrier(5). A growing body of work supports the use of metabolomics to provide novel information about the initiation and progression of PD(5,6). For instance, metabolites related to lipid metabolism, including glycerophospholipids and sphingolipids, mitochondrial function, and amino acids have been implicated in PD(7,8).

Here, we have performed a series of untargeted, agnostic analyses to explore metabolite signatures associated with PD. Using serum-based untargeted, high-performance liquid-chromatography mass spectrometry (LCMS), we performed a hypothesis-generating metabolome-wide association study (MWAS) with independent discovery and replication study populations. We identified individual metabolite features associated with PD, evaluated pathway enrichment, and assessed associations between PD-MWAS metabolites and symptom profiles among patients. Ultimately, identifying disrupted metabolic pathways in PD may improve our understanding of the molecular mechanisms underlying pathogenesis, paving the way for new preventative or therapeutic strategies.

## Methods

### Study Population

We used metabolomics data from 642 PD patients and 277 controls recruited as part of a community-based study of Parkinson’s disease (Parkinson’s Environment and Genes study, PEG). PEG is a population-based PD case-control study conducted in three Central California counties(9). Participants were recruited in two waves: PEG1, 2000-2007 and PEG2, 2011-2018. All those with serum for metabolomics were included (PEG1: n=282 PD patients, n=185 controls; PEG2: n=360 PD patients, n=90 controls). Patients were early in their disease course at enrollment (3.0 years [SD=2.6] on average from diagnosis) and all were seen by UCLA movement disorder specialists for in-person neurologic exams and confirmed as having idiopathic PD based on clinical characteristics(10). Characteristics of the PEG study subjects are shown in Supplemental Table 1. The patients were on average slightly older than the controls and a higher proportion were men, Hispanic, and never smokers.

### High-Resolution Metabolomics (HRM)

HRM profiling was conducted according to established methods. Detailed methods are provided in previous publication(11). Briefly, serum samples were randomly sorted into batches of 40. Each sample was thoroughly mixed with ice-cold acetonitrile (2:1 acetonitrile to serum), placed on ice for 30 minutes, precipitated protein was removed by centrifugation, and the resulting supernatant was transferred to an autosampler vial containing a low volume insert. We analyzed all sample extracts in triplicate with a dual-column, dual-polarity approach, including hydrophilic interaction (HILIC) chromatography with positive electrospray ionization (ESI) and C18 chromatography with negative ESI, and used two types of two quality control samples (see supplement). Details on raw data extraction, feature alignment, and retention time adjustment are also provided in the supplement. Uniquely detected ions consisted of *m*/*z*, retention time (rt), and ion abundance. For analyses, we included metabolomic features with median coefficients of variation among technical replicates <75% and Pearson correlation >0.7 and features detected in >50% of all study samples, leaving 2046 C18 features and 2716 HILIC features for analysis. We log 2 transformed, quantile normalized, and batch corrected the data with ComBat after replacing zeroes with the lowest detected value. Data pre-processing, batch and technical artifact correction, and visualization are further described in the supplemental materials and Supplemental Figures 1-7.

### Metabolome-Wide Association Analysis (MWAS)

To associate features with PD, we conducted unconditional logistic regression for each metabolite with PD as the outcome and age, gender, race/ethnicity, and a year of sample draw indicator included as covariates. We determined metabolite associations independently for the PEG1 and PEG2 study waves and then combined odds ratio (OR) estimates in a fixed effects meta-analysis, using a generic inverse-variance method for pooling(12). We used a false discovery rate (FDR) to correct for multiple testing. Our *a priori* criteria for determining the highest level of PD association were metabolites with a meta-FDR<0.05 and independent association in both discovery (PEG1) and replication (PEG2) populations at p<0.05. For metabolites which showed association with PD, using linear regression, we further tested for association with the following PD and PD symptom related phenotypes among PD patients only: levodopa equivalent daily dose (LEDD), Hoehn Yahr (HY) stage, and Unified Parkinson’s disease Rating Scale Part III (UPDRS-III) score.

### Annotation and Pathway Analysis

We annotated features based on three levels. First, significant features were matched to a database of authenticated chemical standards previously characterized in the Emory laboratory, i.e., metabolites confirmed using MS/MS and authentic standards, providing the strongest level of annotation(13,14). The error tolerance was set to 5 ppm and 30s for *m*/*z* and retention time. Additional *m*/*z* feature mapping was done based on *mummichog* annotations and *xMSannotator. mummichog* is a computational algorithm which uses metabolic pathways and networks to predict functional activity from untargeted metabolite feature tables, including providing annotations of features based upon predicted ions and pathway associations(15). With *xMSannotator*, accurate mass *m*/*z* for adducts formed under positive/negative ESI mode was matched to HMDB, KEGG, and LipidMaps with a mass error threshold of 10 ppm(16). *xMSannotator* uses correlations of intensities and retention time and assigns confidence scores based on a multilevel scoring algorithm (0–3, a higher score representing higher-confidence result), ensuring annotation accuracy. Only results with scores ≥2 were considered for annotations.

For pathway enrichment analysis we used *metapone*, which uses a permutation-based weighted hypergeometric test with joint pathway analysis using positive and negative ion mode data to avoid double counting and account for multiple-matching uncertainty with a weighting factor(17). Metabolic pathways were compiled from KEGG, *mummichog*, and the small molecule pathway database (SMPDB).

## Results

Our untargeted metabolome-wide association study included 4762 features (2046 C18 and 2716 HILIC). We detected 213 associated with PD at a meta-analysis FDR<0.05. Of these features, 104 met the criteria for independent replication (discovery and replication p<0.05). Figure 1A (C18) and 1B (HILIC) show the MWAS results. In total, from the C18 column, 116 features were associated at a meta-analysis FDR<0.05 (372 at p<0.05), 66 of which were independently associated in discovery and replication at p<0.05 (116 at p<0.10). From the HILIC column, 97 features were associated at a meta-analysis FDR<0.05 (441 at p<0.05), 38 of which were associated in discovery and replication at p<0.05 (54 at p<0.10). The full MWAS summary statistics are provided in Supplemental Table 2 (C18 metabolites) and Supplemental Table 3 (HILIC metabolites). Annotation based on three-layers (in-house database of metabolites, *mummichog* annotations, and *xMSannotator* high confidence matches) for all features with discovery and replication at p<0.1 is provided in Supplemental Table 4 (C18) and Supplemental Table 5 (HILIC). The full *xMSannotator* stage 5 annotation results are provided in Supplemental Table 6 (C18) and Supplemental Table 7 (HILIC).

**Figure 1.**
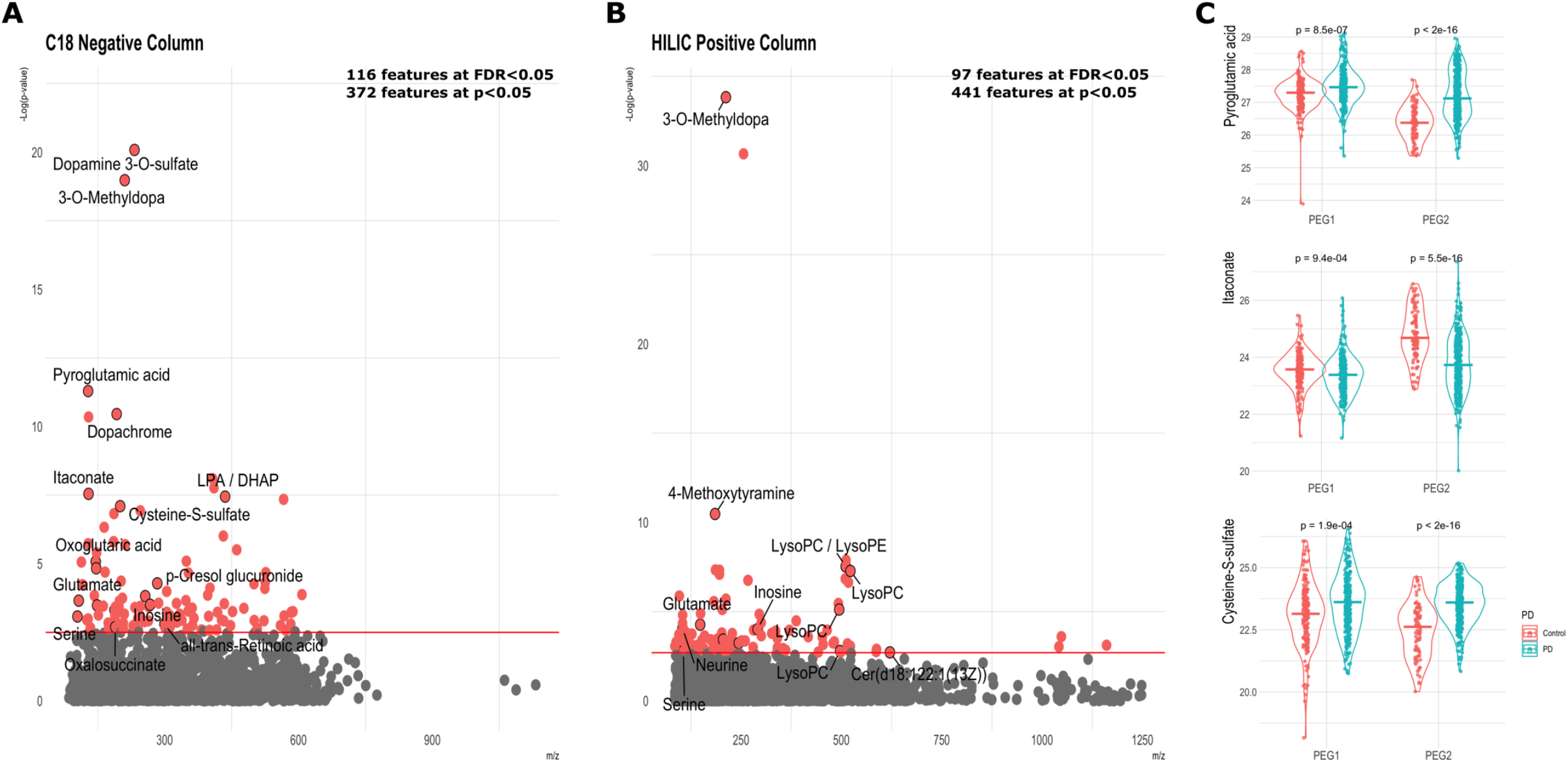
Manhattan Plot of MWAS results from metabolomics LCMS data derived from both the (A) C18 negative ion channel and (B) HILIC positive ion channel. Horizontal lines are shown at FDR≤0.05. (C) Violin plots show the top three annotated metabolites from the MWAS, excluding PD medication-associated metabolites, and separated by independent study populations.

Table 1 shows the top MWAS metabolites that were associated with PD in discovery and replication. As expected, the leading PD-associated features from both columns were related to PD-medications, including medication metabolites, dopamine 3-O-sulfate and 3-O-methyldopa. In total, 11 C18 features and 16 HILIC features were associated with LEDD at an FDR<0.05 among the PD patients (Supplemental Tables 2 and 3). Additional metabolites strongly associated with PD included pyroglutamic acid (meta-OR= 3.79, 95% CI=2.6, 5.54, FDR=3.5e-9), itaconate (meta-OR= 0.47, 95% CI=0.36, 0.61, FDR=7.3e-6), and cysteine-S-sulfate (meta-OR= 1.56, 95% CI=1.32, 1.83, FDR=8.2e-6) from the C18 column (Figure 1C). In the HILIC column, a series of PD distinguishing features annotated to multiple phospholipids, including lysophosphatidylcholines (LysoPC) and lysophosphatidylethanolamines (LysoPE). A LysoPC(18:1), for example, was found at higher intensity among patients in both discovery and replication (meta-OR= 3.85, 95% CI=2.37, 6.27, FDR=1.8e-5). Features from both columns which annotated to inosine were inversely associated with PD (C18: meta-OR= 0.82, 95% CI=0.74, 0.92, FDR=1.4e-2; HLIIC: meta-OR= 0.91, 95% CI=0.87, 0.96, FDR=9.4e-3).

**Table 1.** PD MWAS hits: Annotated features from the MWAS associated with Parkinson’s disease in both discovery and replication study populations.

| mz | time | Meta OR<br>(95% CI) | Meta P | Meta<br>FDR | Discovery<br>(PEG1) | Replication<br>(PEG2) | LEDD p-<br>value | LEDD FDR | Compound Annotation* | Number of<br>xmsAnnotator<br>matches (≥2) |
| --- | --- | --- | --- | --- | --- | --- | --- | --- | --- | --- |
| C18 Negative Ion Column |  |  |  |  |  |  |  |  |  |  |
| 232.0282 | 34.8165 | 1.24 (1.18, 1.29) | 8.35E-21 | 1.71E-17 | 1.05E-16 | 1.15E-05 | 8.70E-37 | 8.90E-34 | Dopamine 3-O-sulfate<sup>2,3</sup> | 2 |
| 210.0772 | 35.9778 | 1.72 (1.53, 1.94) | 1.06E-19 | 1.09E-16 | 4.49E-15 | 2.33E-07 | 3.17E-76 | 6.48E-73 | 3-O-Methyldopa<sup>1,2,3</sup> | 3 |
| 128.0353 | 30.6303 | 3.79 (2.60, 5.54) | 5.14E-12 | 3.51E-09 | 4.86E-07 | 1.42E-08 | 1.84E-01 | 7.09E-01 | Pyroglutamic acid/<br>Pyrrolidonecarboxylic acid<sup>1,3</sup> | 7 |
| 192.0305 | 24.1011 | 1.55 (1.36, 1.76) | 3.54E-11 | 1.81E-08 | 7.84E-10 | 4.42E-03 | 2.44E-26 | 1.67E-23 | Dopachrome<sup>2,3</sup> | 3 |
| 129.0192 | 33.8177 | 0.47 (0.36, 0.61) | 2.86E-08 | 7.32E-06 | 6.14E-04 | 9.36E-07 | 5.10E-02 | 5.24E-01 | Itaconate (Itaconic acid)<sup>1,3</sup> | 9 |
| 435.2514 | 167.2708 | 1.62 (1.36, 1.92) | 3.61E-08 | 8.20E-06 | 1.01E-04 | 8.85E-05 | 1.09E-02 | 3.42E-01 | LPA(0:018:1(9Z));<br>LPA(18:1(9Z)0:0); DHAP(18:0)<sup>3</sup> | 3 |
| 199.9693 | 28.4295 | 1.56 (1.32, 1.83) | 8.03E-08 | 1.49E-05 | 7.10E-05 | 2.95E-10 | 1.29E-01 | 6.33E-01 | Cysteine-S-sulfate<sup>3</sup> | 1 |
| 145.0143 | 28.1639 | 1.61 (1.31, 1.98) | 8.38E-06 | 7.78E-04 | 9.43E-04 | 2.79E-04 | 5.46E-02 | 5.32E-01 | Oxoglutaric acid<sup>1,2,3</sup> | 1 |
| 146.0459 | 29.4772 | 1.86 (1.40, 2.46) | 1.48E-05 | 1.26E-03 | 2.98E-03 | 4.24E-04 | 7.60E-02 | 5.94E-01 | Glutamic acid<sup>2,3</sup> | 1 |
| 283.0821 | 30.1765 | 1.10 (1.05, 1.15) | 5.22E-05 | 3.56E-03 | 3.63E-04 | 5.57E-02 | 8.07E-01 | 9.66E-01 | p-Cresol glucuronide<sup>3</sup> | 3 |
| 256.0936 | 28.4837 | 1.14 (1.06, 1.22) | 1.52E-04 | 7.97E-03 | 4.35E-03 | 2.03E-05 | 7.59E-02 | 5.94E-01 | Glycerophosphocholine<sup>3</sup> | 3 |
| 107.0502 | 34.5396 | 1.29 (1.13, 1.47) | 2.23E-04 | 1.08E-02 | 5.43E-03 | 7.76E-03 | 2.44E-01 | 7.52E-01 | p-Cresol<sup>3</sup> | 5 |
| 267.0723 | 32.8807 | 0.82 (0.74, 0.92) | 3.13E-04 | 1.36E-02 | 1.67E-02 | 3.13E-05 | 2.85E-02 | 4.51E-01 | Inosine<sup>2,3</sup> | 4 |
| 148.0405 | 31.5075 | 1.27 (1.12, 1.45) | 3.31E-04 | 1.41E-02 | 1.12E-02 | 2.23E-03 | 4.38E-20 | 1.49E-17 | 5,6-Dihydroxyindole<sup>2,3</sup> | 3 |
| 187.0071 | 35.7939 | 1.24 (1.10, 1.39) | 4.94E-04 | 1.77E-02 | 1.03E-02 | 1.06E-02 | 3.46E-01 | 8.40E-01 | p-Cresol sulfate<sup>3</sup> | 2 |
| 151.0514 | 33.6125 | 0.84 (0.77, 0.93) | 7.45E-04 | 2.21E-02 | 8.23E-03 | 2.32E-02 | 5.50E-01 | 9.03E-01 | N1-Methyl-4-pyridone-5-carboxamide<sup>3</sup> | 2 |
| 104.0353 | 30.1006 | 2.20 (1.39, 3.51) | 8.43E-04 | 2.27E-02 | 3.48E-02 | 1.98E-03 | 3.76E-03 | 2.31E-01 | Serine<sup>1,2,3</sup> | 1 |
| 172.9914 | 34.1845 | 0.83 (0.74, 0.93) | 1.46E-03 | 3.21E-02 | 4.29E-02 | 4.95E-03 | 3.51E-01 | 8.42E-01 | Phenol sulphate<sup>3</sup> | 1 |
| 299.2014 | 200.4142 | 1.16 (1.06, 1.26) | 1.60E-03 | 3.34E-02 | 8.45E-02 | 1.82E-04 | 1.21E-02 | 3.51E-01 | all-trans-Retinoic acid<sup>1,3</sup> | 12 |
| 189.0029 | 34.3209 | 1.17 (1.06, 1.29) | 2.04E-03 | 3.98E-02 | 3.58E-02 | 1.26E-02 | 2.64E-01 | 7.82E-01 | Oxalosuccinate<sup>2,3</sup> | 1 |
| HILIC Positive Ion Column |  |  |  |  |  |  |  |  |  |  |
| 212.0917 | 60.9225 | 1.43 (1.35, 1.52) | 1.49E-34 | 4.04E-31 | 1.08E-28 | 5.49E-08 | 3.32E-70 | 9.03E-67 | 3-O-Methyldopa<sup>3</sup> | 3 |
| 185.1285 | 46.3928 | 0.42 (0.32, 0.54) | 3.37E-11 | 3.05E-08 | 1.34E-07 | 3.79E-05 | 2.37E-07 | 1.61E-04 | 4-Methoxytyramine<sup>3</sup> | 0 |
| 510.3557 | 47.8290 | 4.06 (2.47, 6.65) | 2.92E-08 | 1.59E-05 | 8.16E-04 | 1.52E-08 | 1.30E-02 | 2.78E-01 | LysoPC(17:0); LysoPE(0:020:0);<br>LysoPE(20:00:0)<sup>3</sup> | 3 |
| 522.3553 | 48.2820 | 3.85 (2.37, 6.27) | 5.27E-08 | 1.79E-05 | 3.98E-03 | 5.78E-09 | 8.25E-03 | 2.49E-01 | LysoPC(18:1(9Z));<br>LysoPC(18:1(11Z))<sup>3</sup> | 2 |
| 494.3237 | 49.3839 | 2.41 (1.64, 3.54) | 7.69E-06 | 1.16E-03 | 1.04E-02 | 4.02E-06 | 9.77E-01 | 9.98E-01 | LysoPC(16:1(9Z))<sup>3</sup> | 1 |
| 149.0597 | 60.1071 | 3.05 (1.85, 5.03) | 1.28E-05 | 1.83E-03 | 4.75E-02 | 1.77E-09 | 5.37E-19 | 4.87E-16 | 3-Methoxy-4-hydroxyphenylacetaldehyde<sup>2</sup> | 7 |
| 148.0604 | 89.4789 | 1.63 (1.29, 2.08) | 5.41E-05 | 5.88E-03 | 1.10E-02 | 8.96E-05 | 1.71E-01 | 6.41E-01 | L-Glutamate<sup>1,3</sup> | 8 |
| 104.1070 | 54.8209 | 3.11 (1.77, 5.47) | 8.24E-05 | 8.61E-03 | 8.75E-02 | 3.06E-05 | 5.12E-02 | 4.16E-01 | L-Isoleucine<sup>2</sup>; Neurine<sup>3</sup> | 1 |
| 291.0693 | 63.1794 | 0.91 (0.87, 0.96) | 9.66E-05 | 9.37E-03 | 2.49E-02 | 6.93E-09 | 2.01E-01 | 6.68E-01 | Inosine<sup>2</sup> | 0 |
| 205.0972 | 59.8331 | 0.46 (0.30, 0.71) | 3.70E-04 | 2.06E-02 | 3.15E-03 | 4.40E-02 | 7.25E-01 | 9.39E-01 | D-Tryptophan<sup>2,3</sup> | 8 |
| 244.1542 | 37.5565 | 0.75 (0.64, 0.89) | 5.77E-04 | 2.62E-02 | 7.84E-03 | 2.02E-02 | 7.77E-01 | 9.50E-01 | Tiglylcarnitine<sup>3</sup> | 1 |
| 106.0499 | 98.4517 | 1.90 (1.28, 2.83) | 1.59E-03 | 4.64E-02 | 3.30E-02 | 1.01E-02 | 2.29E-02 | 3.34E-01 | Serine<sup>1,2,3</sup> | 2 |
| 496.3396 | 49.1793 | 2.33 (1.38, 3.93) | 1.61E-03 | 4.65E-02 | 7.22E-02 | 5.63E-03 | 2.90E-01 | 7.58E-01 | LysoPC(16:0)<sup>3</sup> | 1 |
| 620.5915 | 68.3020 | 1.46 (1.15, 1.86) | 1.92E-03 | 5.05E-02 | 5.77E-02 | 1.62E-03 | 2.66E-01 | 7.40E-01 | Cer(d18:122:1(13Z))<sup>3</sup> | 1 |

\*Annotation: 1: In House Library, 2: Mummichog Best Guess, 3: xmsAnnotator High Confidence (level 2 or 3 confidence)

P-cresol and two of its metabolites, p-cresol sulfate and p-cresol glucuronide, were also found at higher intensity among the PD patients relative to controls (p-cresol meta-OR=1.29, 95% CI=1.13, 1.47, FDR=0.01). The distributions of these metabolites by PD and across discovery and replication populations are shown in Figure 2A. The p-cresol metabolites were also correlated with age at PD diagnosis (Figure 2B) and p-cresol glucuronide was associated with a higher Hoehn Yahr (HY) stage among PD patients (beta=0.02, SE=0.007, FDR=9.5e-2; Figure 2C). In fact, of all PD-associated features (MWAS meta-p<0.05), seven were also related to HY stage among PD patients at an FDR<0.05 (115 metabolites associated at p<0.05), including, as expected, the PD medication metabolite, 3-O-methyldopa, providing confidence in the results (Supplemental Table 8). Six PD-associated features were also associated with UPDRS-III at an FDR<0.05 (100 metabolites at p<0.05; Supplemental Table 9). However, other than the PD medication metabolites and p-cresol glucuronide, the features associated with either HY stage or UPDRS-III up to FDR<0.10 could not be annotated at high confidence.

**Figure 2.**
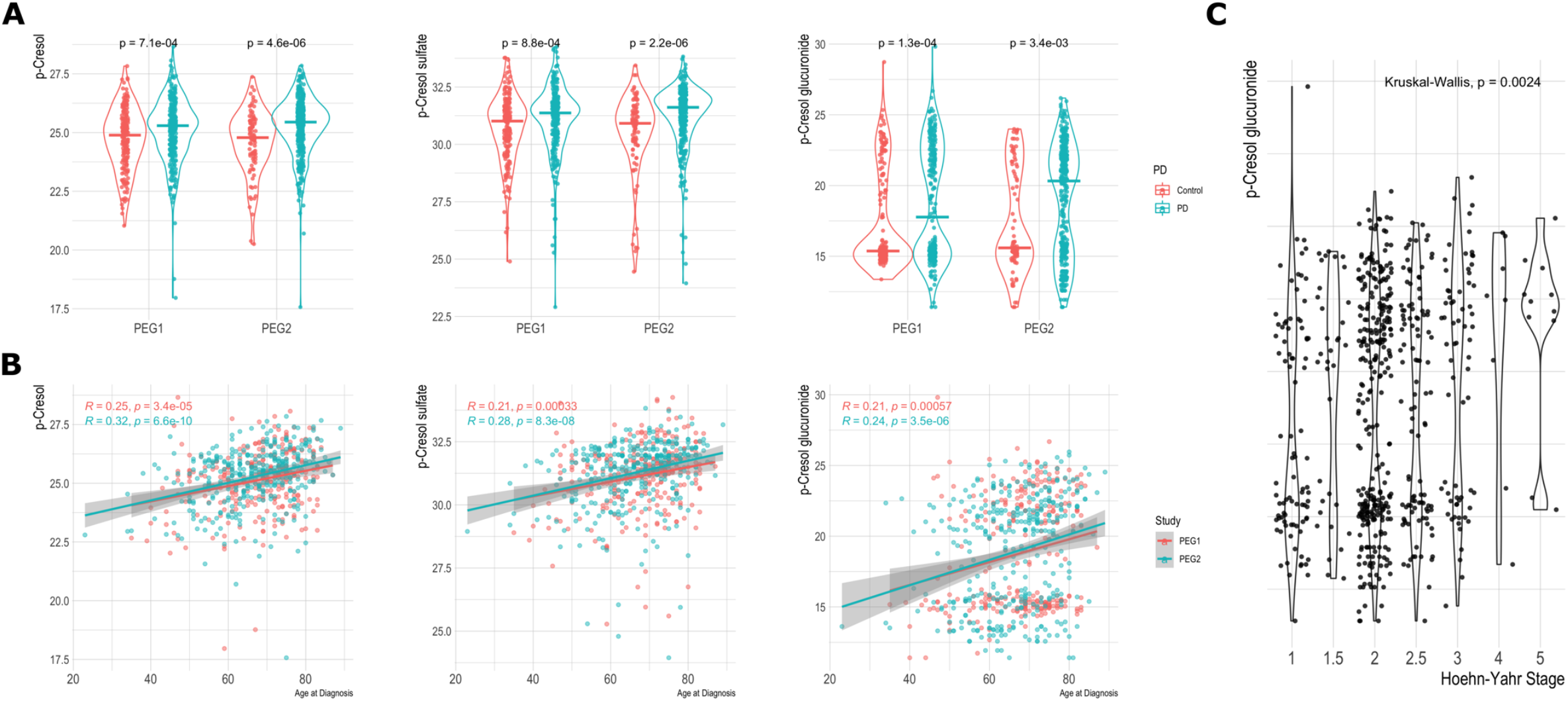
P-cresol and two p-cresol metabolites are associated with (A) Parkinson’s disease, (B) age at diagnosis among PD patients, and (C) Hoehn- Yahr Stage among PD patients.

Given the interdependent nature of metabolites, we further assessed correlation patterns between the PD-associated features. Figure 3 shows a Pearson-correlation based network of all FDR<0.05 MWAS features from both columns. Several highly correlated clusters of features are visible, including a PD medication related cluster, a phospholipids cluster, and a cluster of several features correlated with p-cresol.

**Figure 3.**
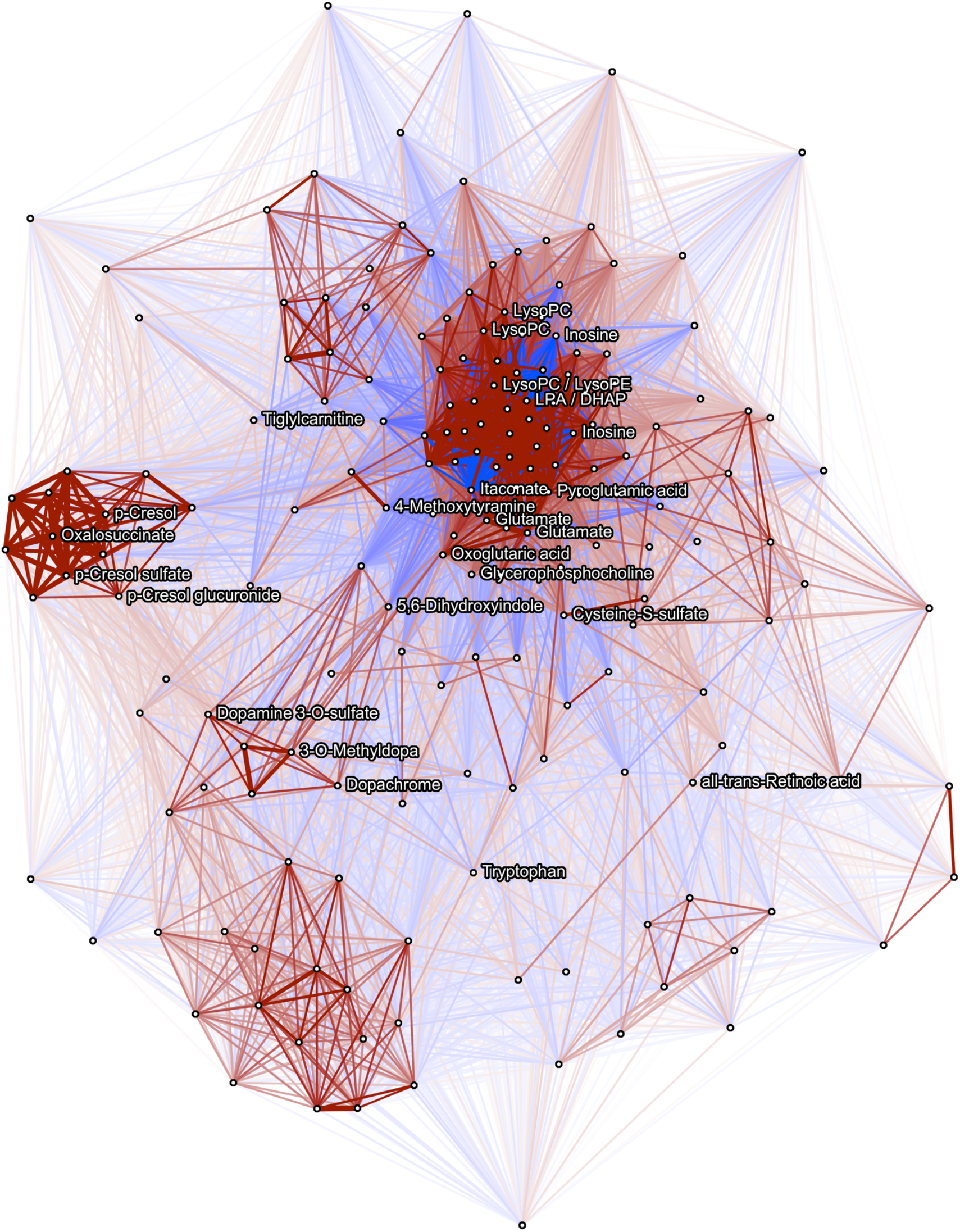
Pearson correlation network between MWAS FDR<0.05 metabolites from both the C18 and HILIC columns. |Correlations|≥0.2 shown. Features which were annotated are named to the right, while other features are shown as blank nodes.

Multiple pathways were also overrepresented in the MWAS features. Based on metabolic pathway analysis, 17 pathways were overrepresented at an FDR<0.05 (58 at p<0.05; Figure 4 and Supplemental Table 10). Glutamine and glutamate metabolism, gabaergic synapse, methionine and cysteine metabolism, glycine, serine, alanine and threonine metabolism, and leukotriene metabolism were among the most significantly overrepresented pathways. Several phospholipid pathways, including glycerophospholipid and glycosphingolipid metabolism and phospholipase d and sphingolipid signaling pathways, were also overrepresented among the PD-associated metabolites.

**Figure 4.**
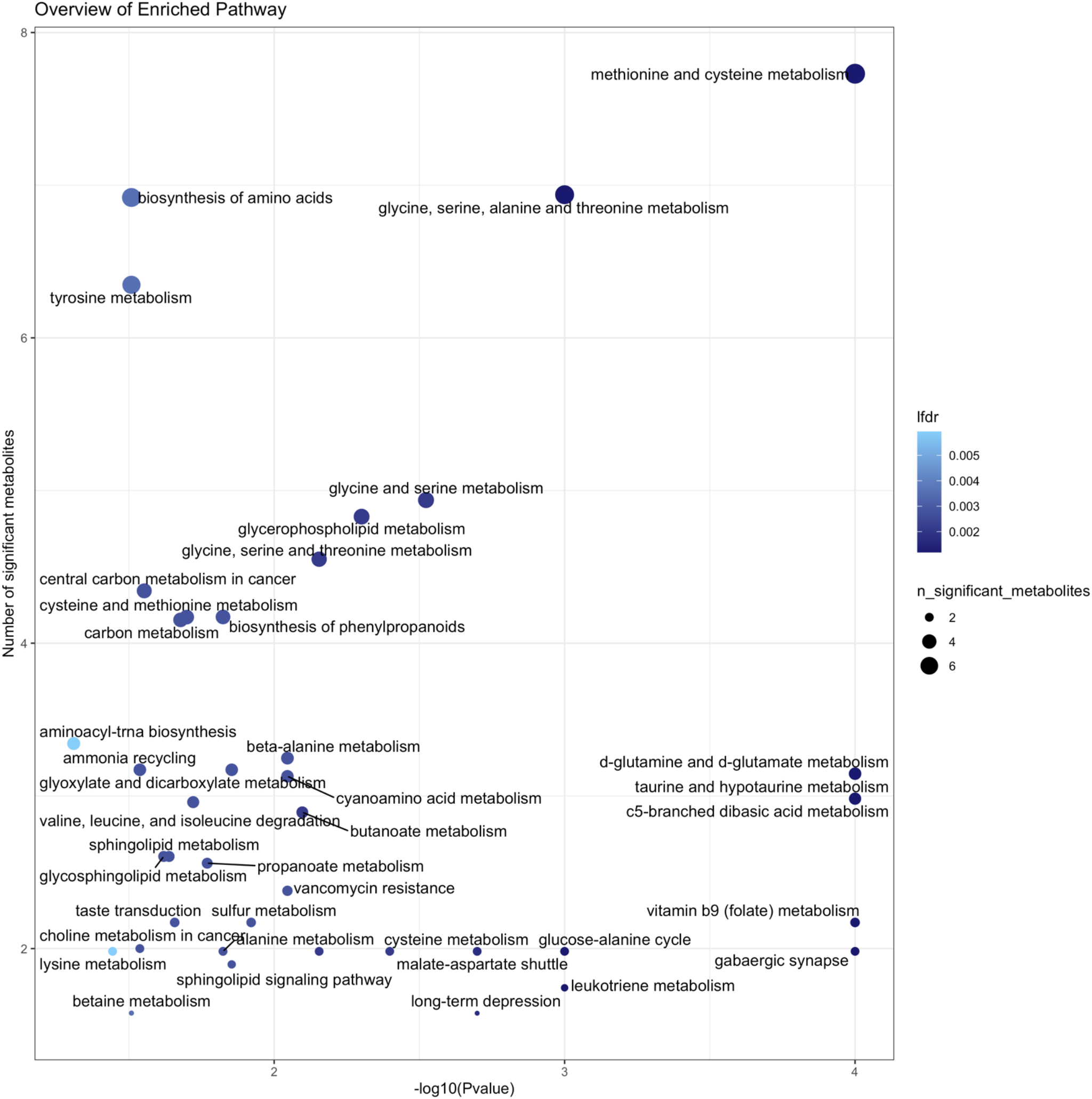
Overview of enriched pathway analysis. Based on pathway analysis of untargeted PD MWAS features using a permutation-based weighted hypergeometric test (R, *Metapone: a Bioconductor package for joint pathway testing for untargeted metabolomics data*). Pathways with p<0.05 are shown. Lfdr= the local FDR value for each enrichment.

## Discussion

Using high resolution, untargeted serum metabolic profiling based on LC-MS, we identified 213 metabolite features associated with PD and 17 overrepresented metabolic pathways. Our agnostic metabolomics approach implicated amino acid metabolism and phospholipid pathways as important in PD, along with multiple individual metabolites with compelling links to neurodegeneration. Serum was collected from PD patients early in disease and compared to community controls. These metabolites and pathways therefore may reflect disturbances due to disease pathogenesis and progression as well as compensatory or reactive mechanisms caused by disease or treatment.

One of the strengths of our study is that we assessed metabolite profiles from a large number of PD patients recruited early in disease who were undergoing a range of treatment courses. Thus, we were able to assess the relationship between all metabolite features and levodopa equivalent daily dose to determine which features associated with PD were also associated with medication use. Predictably, PD patients differed from controls most strongly in terms of levodopa metabolites or metabolites involved in dopamine metabolism. Network analysis further showed the PD medication metabolites clustered together, as expected, but were not significantly correlated with other PD-related metabolites. Several other pathways and specific metabolites were implicated independent of levodopa medication use. Another noteworthy metabolite that attests to the validity of our analyses is inosine, which is a precursor to urate, with anti-inflammatory properties. We observed inosine at lower intensity among the patients relative to controls. This is in line with previous studies and the notion that lower uric acid level may be involved in faster PD progression, which have even resulted in inosine supplementation trials(18–21).

One of the most intriguing findings from this analysis was p-cresol and its two metabolites, p-cresol sulfate and p-cresol glucuronide. We observed higher intensities of the metabolites among PD patients relative to controls in both discovery and replication populations. Moreover, among patients, intensity of all three p-cresol metabolites was related to age at diagnosis, with older patients showing higher intensity and p-cresol glucuronide was related to higher motor symptom scores. P-cresol is an exogenous uremic toxin primarily produced by gut bacteria which express p-cresol synthesizing enzymes that are not produced by human cells. Interestingly, while this is the first study to implicate these metabolites in blood, a small study previously found higher levels of p-cresol in the CSF of PD patients(22). Multiple studies have also linked p-cresol with autism(23– 25) and altered brain dopamine metabolism in neurodevelopment(26). Furthermore, gut dysbiosis has been linked to both PD and autism, including among our own patients(27), with some research even indicating that misfolded a-synuclein retrogradely propagates from the enteric to the central nervous system(28). P-cresol and its’ metabolites therefore represent very compelling targets for future mechanistic research.

Several tricarboxylic acid (TCA) cycle metabolites were also implicated as relevant to PD in our MWAS. PD patients had higher relative abundance of oxoglutaric acid (e.g. alpha-ketoglutarate) and lower levels of itaconate. Aside from clear implications for energy metabolism, which is highly pertinent given the known mitochondrial dysfunction in PD, itaconate holds key roles in immunometabolism (e.g., changes of metabolic pathways within immune cells)(29,30). Itaconate is a mitochondrial metabolite, produced in high amounts by macrophages and monocytes by diverting aconitate away from the TCA cycle during inflammatory activation(30). The primary function appears to be anti-inflammatory, supported by human studies showing that low levels of plasma itaconate coincide with excessive inflammation(30). Inflammation and neuroinflammation are principal features of PD. Thus, it is quite interesting that in both of our patient populations we found lower relative levels of this important anti-inflammatory immunometabolite.

We further identified several amino acids as differentially abundant in PD. Glutamate and several connected metabolites, including pyroglutamic acid (PGA), had a higher relative abundance in patients’ serum relative to controls, with the glutamine and glutamate metabolism pathways significantly overrepresented. PGA is an endogenous metabolite derived from glutamate and linked to glutathione turnover(31,32). Elevated serum PGA therefore may be related to perturbed glutathione metabolism. Low levels of the antioxidant glutathione are an early neuronal biochemical finding in PD(33). But increased systemic levels of PGA may reflect an upregulation of glutathione metabolism to counter inflammatory states and oxidative stress in PD. Furthermore, the neurotransmitter glutamate itself has been linked to PD pathogenesis, with several, though not all, studies reporting increased blood-measured levels of glutamate(34,35). Other amino acid metabolic pathways, including methionine and cysteine metabolism, glycine, serine, alanine and threonine metabolism, and valine, leucine, and isoleucine degradation were also overrepresented among PD-associated features, with individual metabolites like serine, isoleucine, and tryptophan observed in higher relative abundance among the PD patients. The patients also had higher levels of cysteine-S-sulfate, a purportedly brain damaging metabolite involved in sulfite oxidase deficiency(36). Branched chain amino acids (BCAAs), including leucine, isoleucine, and valine, have been linked to PD before, as BCAAs are involved in energy metabolism, preventing oxidative damage, and regulation of protein synthesis(5).

Lipid pathways and metabolites were also strongly implicated with PD by our MWAS. Glycerophospholipid along with glycosphingolipid and sphingolipid metabolism were enriched in pathway analyses. Metabolites including glycerophosphocholine, several lysophosphotidylcholines (LysoPC), and a ceramide, were all observed at higher intensities among the patients relative to controls. The lipid profile in PD has received a great deal of interest in recent years, in part due the identification of *GBA* variants in PD GWAS. The glucosylceramidase-beta (*GBA*) gene, which encodes the lysosomal enzyme glucocerebrosidase (GCase), has directly connected lipid and sphingolipid metabolism to PD pathogenesis(37). PD pathogenic mechanisms linked to lipid metabolism include oxidative stress, inflammation and immune system signaling, pro-apoptotic processes, and interaction with a-synuclein biology, among others. Furthermore, alterations in serum, plasma, and brain measured phospholipids and sphingolipids have been widely reported in PD(38). For instance, LysoPC(18:1), implicated in our MWAS, has also been found at higher levels in the substantia nigra in animal models of PD(39). Interestingly, Cer(d18) metabolites, one of which was observed at higher intensity among our PD patients, have also been associated with physical frailty among older adults(40).

Our study is the largest untargeted high-resolution metabolomics study of PD to date, with metabolic profiles from independent discovery and replication case-control study populations allowing for validation of associated features. However, a notable limitation of the untargeted LC-MS technology is feature annotation. High-resolution LC-MS metabolomics provide metabolite features, many of which are not identified and can only be annotated based on m/z and retention time parameters from large databases (e.g. HMDB) and with consideration of feature correlation structures. While this does allow high confidence annotation, future research will be needed to identify features with certainty. For instance, para-, ortho-, and meta-cresol are all isomers. We have labeled the cresol metabolite as p-cresol due to co-occurrence with the p-cresol metabolites p-cresol sulfate and p-cresol glucuronide and because of the three exogenous metabolites, it is produced in humans via gut microbes. However, future studies will be needed to resolve the isomers. Furthermore, one-to-many matching and no matching add further uncertainty to feature annotation. Many of the features associated with PD in our MWAS, including some of the most significantly associated features, could not be annotated. Additionally, the metabolome measurements were based on a single blood-draw. Future longitudinal studies will be very informative in disentangling which if any metabolites implicated here are causally related to PD versus disease progression or reactive mechanisms.

In conclusion, based on untargeted high resolution, serum metabolic profiling from LC-MS, we have implicated over 200 individual metabolite features in PD along with multiple metabolic pathways. Several hits implicated pathways known to be disrupted in PD, including amino acid and lipid metabolism. We also present many novel findings, including for itaconate, connecting impaired anti-inflammatory signaling through immunometabolism, and three p-cresol metabolites, linking gut microbial activity to PD.

## Data Availability

Data produced in the present study are available upon reasonable request to the authors

