## Supplemental Materials for "Untargeted serum metabolomics reveals novel metabolite associations and disruptions in amino acid and lipid metabolism in Parkinson’s disease"

**Sample Collection**

Blood samples were drawn from participants during field visits. Samples were centrifuged, kept on dry ice, and then stored in a −80 °C freezer at UCLA. Serum samples were shipped frozen to Emory University on dry ice for metabolomics analyses, where they were stored at −80 °C until analyses.

**High-Resolution Metabolomics**

We included two methods of performance quality control. First, a NIST 1950 QC sample was analyzed at the beginning and end of the entire analytical run(1). A second QC sample (Q-Std), which is commercially purchased plasma pooled from an unknown number of males and females, was analyzed at the beginning, middle, and end of each batch of 40 samples for normalization and batch effect evaluation.

Our samples were processed across two LCMS runs conducted approximately 6-months apart, to pool the metabolite data across runs, we used the *apLCMS* R package to perform retention time adjustment and feature alignment separately for both HILIC and C18 feature tables, using the adjust.time and feature.align functions. For feature alignment, the *m/z* tolerance was 1e-05 and retention time tolerance was 37.016 (C18) and 38.246 (HILIC) seconds. Overall, 2226 features aligned for C18 and 2919 for HILIC across the two LCMS runs. We included metabolomic features detected in >50% of all study serum samples, leaving 2046 C18 features and 2716 HILIC features for analysis.

We log 2 transformed the metabolite data, quantile normalized, and batch corrected with ComBat after replacing zeroes with the lowest detected value which has been recommended for metabolomics data. Data pre-processing and visualization is described and shown in detail in the supplemental materials and Supplemental Figures 1-4. From principal component (PC) analysis with the HILIC features, we discovered two clear clusters of samples seemingly separating based on technical, non-biologic factors. As a result, we performed an additional correction to remove variation between the PCs (Supplemental Figures 5-7).


**1.** Simón-Manso Y, Lowenthal MS, Kilpatrick LE, Sampson ML, Telu KH, Rudnick PA, et al. Metabolite profiling of a NIST standard reference material for human plasma (SRM 1950): GC-MS, LC-MS, NMR, and clinical laboratory analyses, libraries, and

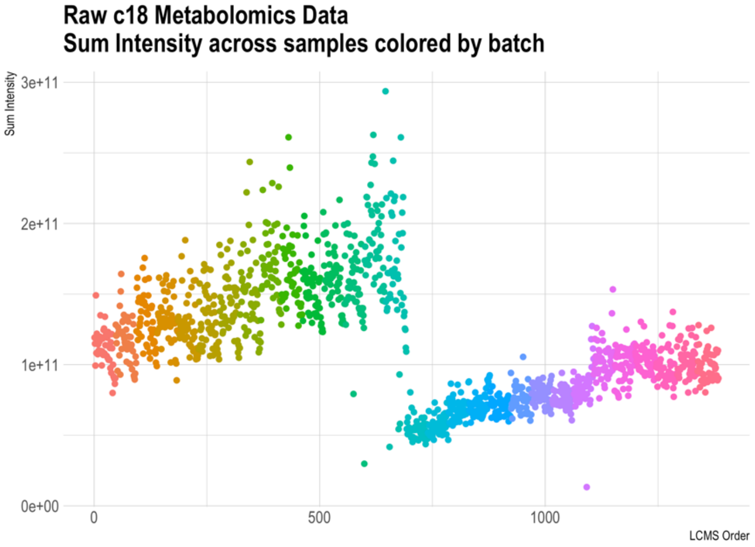


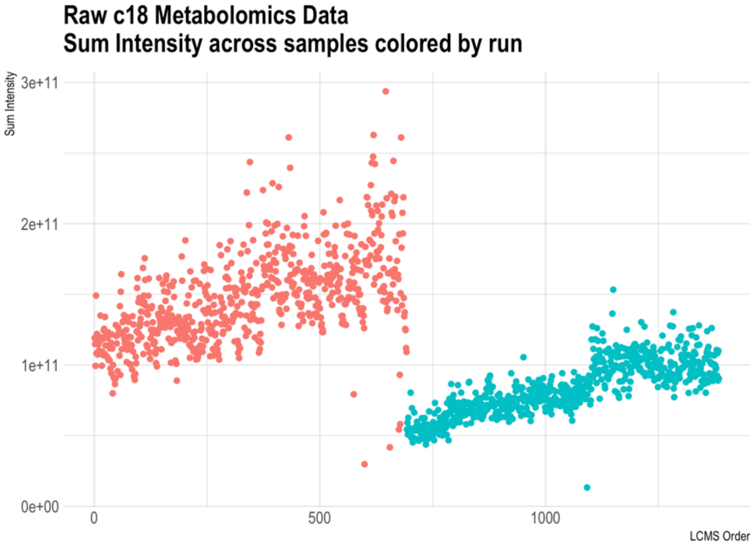


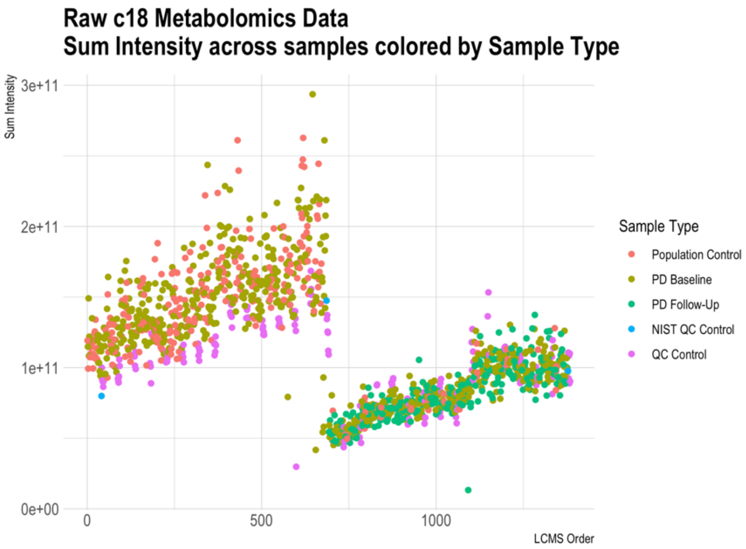


**Supplemental Figure 1. C18 negative column metabolomics processing:** Sum of metabolite intensities across samples colored by batch & sample type, before pre-processing (log transformation, quantile normalization, ComBat batch correction). LCMS was run across 30 batches (n=46); machine was reset after 694 samples (i.e., samples ran in two larger groups of n=694 samples, each with 15 smaller batches within run). Run, batch, and drift effects are apparent in raw data.

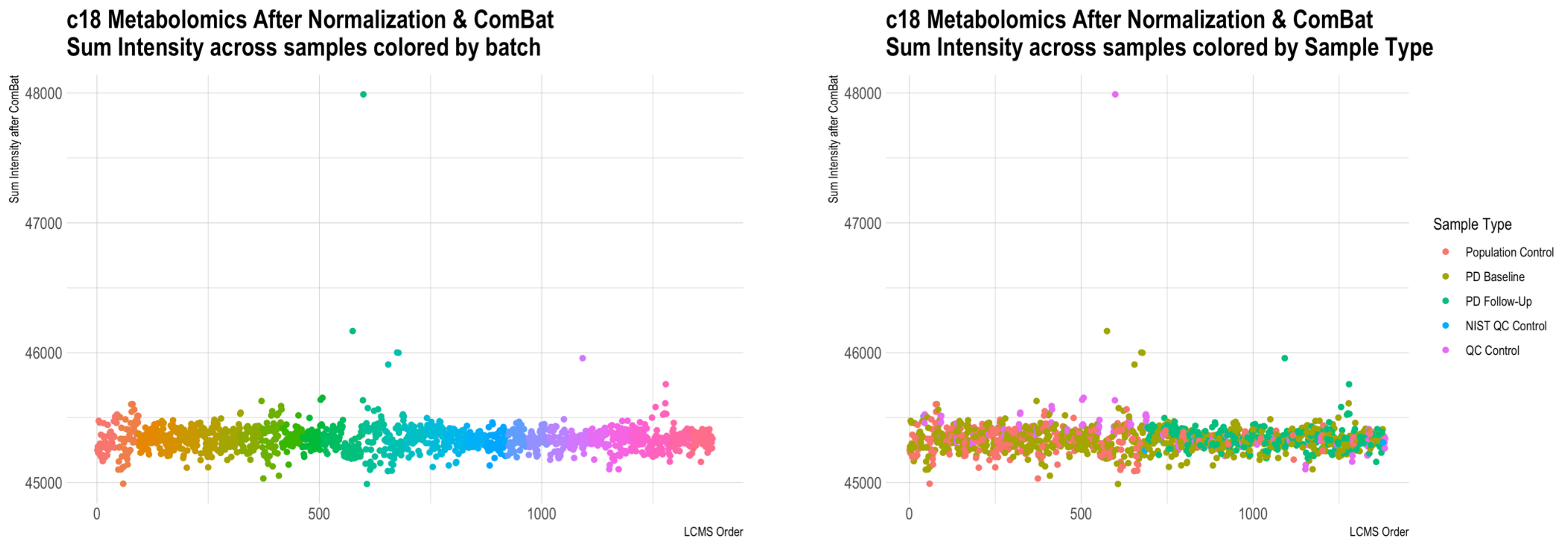


**Supplemental Figure 2. C18 negative column after metabolomics processing.** Raw c18 data was log transformation, quantile normalized, followed by ComBat for batch correction. LCMS was run across 30 batches (n=46); machine was reset after 694 samples (i.e., samples ran in two larger groups of n=694 samples, each with 15 smaller batches within run. While there are several apparent outliers, after processing, technical variation has been removed.

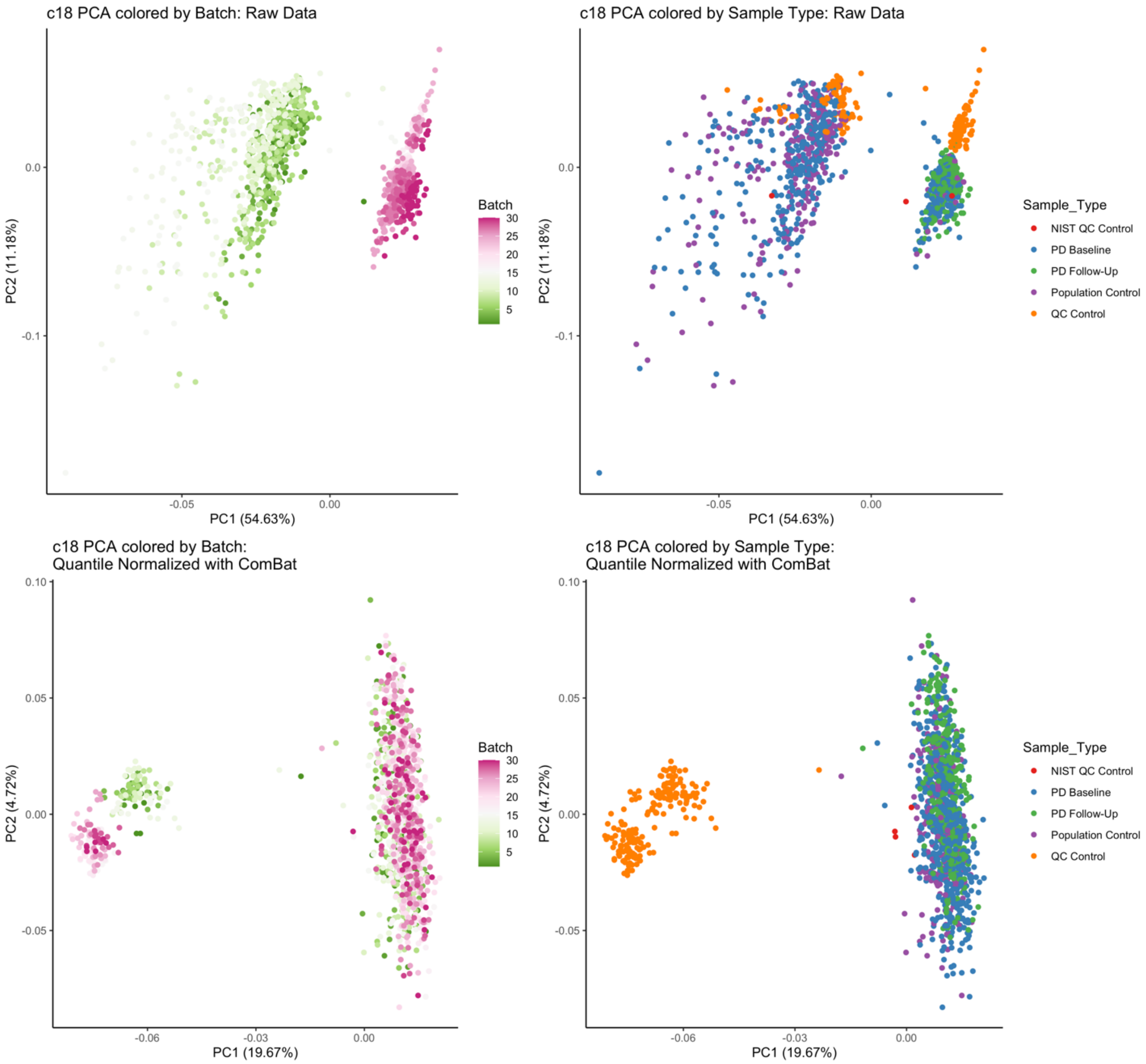


**Supplemental Figure 3. C18 negative column metabolomics processing:** Principal component analysis of raw and processed metabolomics data. PC variation primarily explained by LCMS run in raw data. After correction, sample type (quality control sample versus the study serum samples) primarily explains variation.

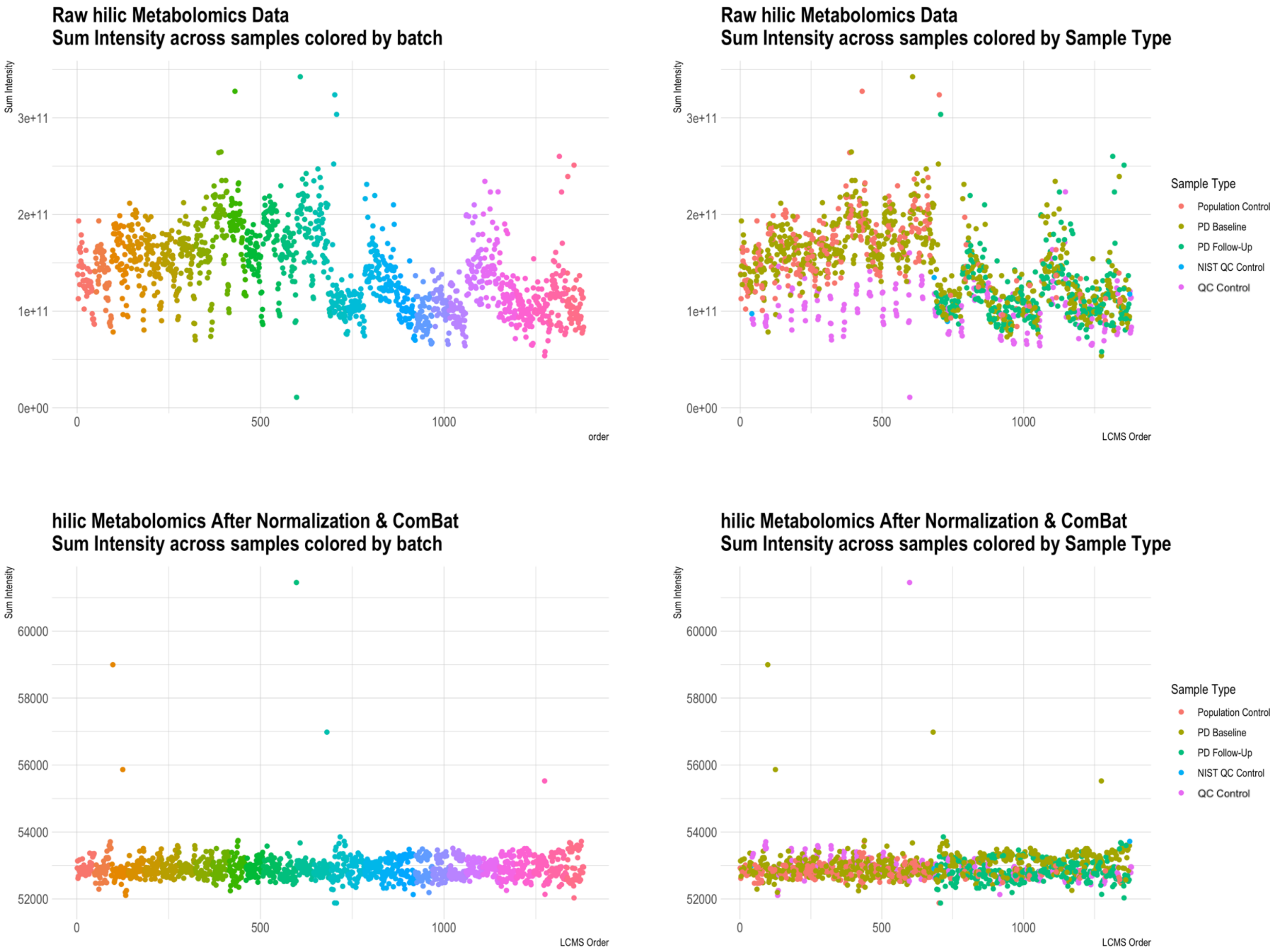


**Supplemental Figure 4. HILIC positive column metabolomics processing:** Sum of metabolite intensities across samples colored by batch & sample type before and after pre-processing (log transformation, quantile normalization, ComBat batch correction). LCMS ran in across 30 batches (n=46);

machine was reset after 694 samples (i.e., samples ran in two larger groups of n=694 samples, each with 15 smaller batches within run). Run, batch, and drift effects are apparent in raw data. While there are several apparent outliers, after processing, the technical variation has been removed.

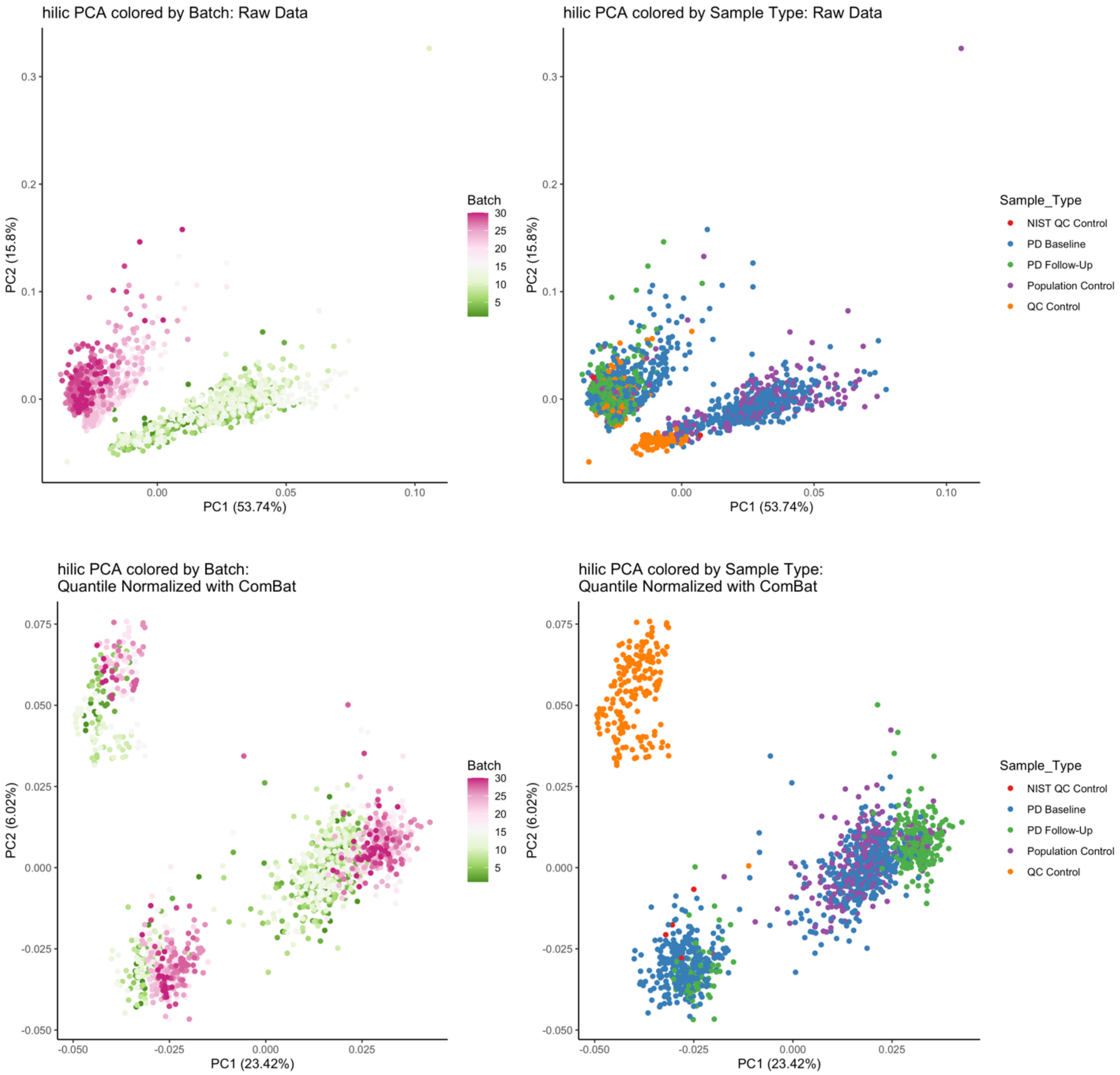


**Supplemental Figure 5. HILIC positive column metabolomics processing:** Principal component analysis of metabolomics data after median normalization and ComBat correction for batch effects. PC variation primarily explained by batch in raw data, after correction sample type (quality control sample versus the population-based serum samples) primarily explains variation. However, there are two apparent clusters of population-based serum samples, potentially explained by non-biologic (PD) technical variation (see **Supplemental Figure 6**).

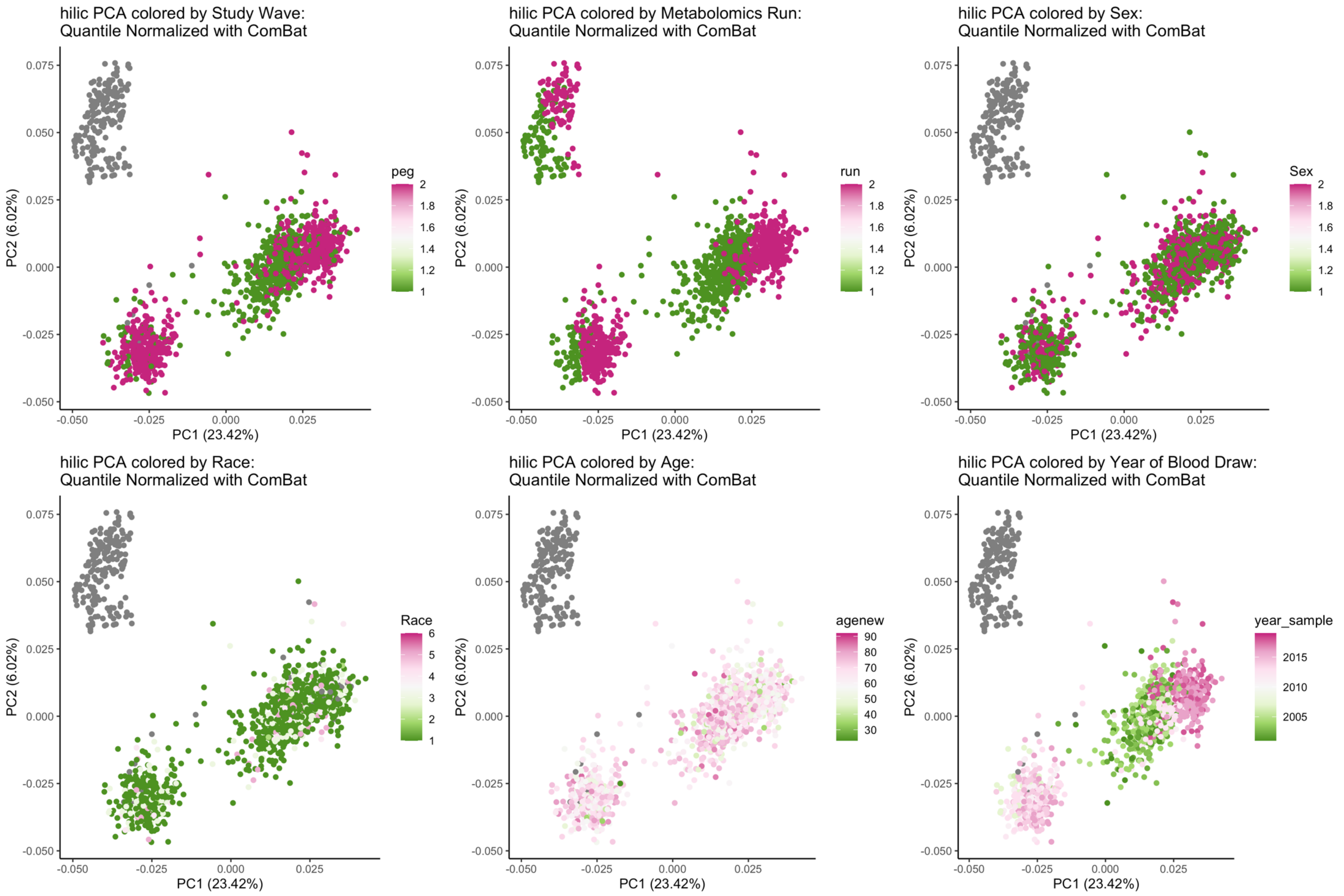


**Supplemental Figure 6. HILIC positive PCA of processed data, colored by different covariates.** No completely distinguishing variables to describe the different clusters of study samples, though there is some separation by year of sample. Note gray indicates the QC samples. Therefore, we additionally corrected for inclusion in this cluster, as variation appears technical and is very influential (Supplemental Figure 7).

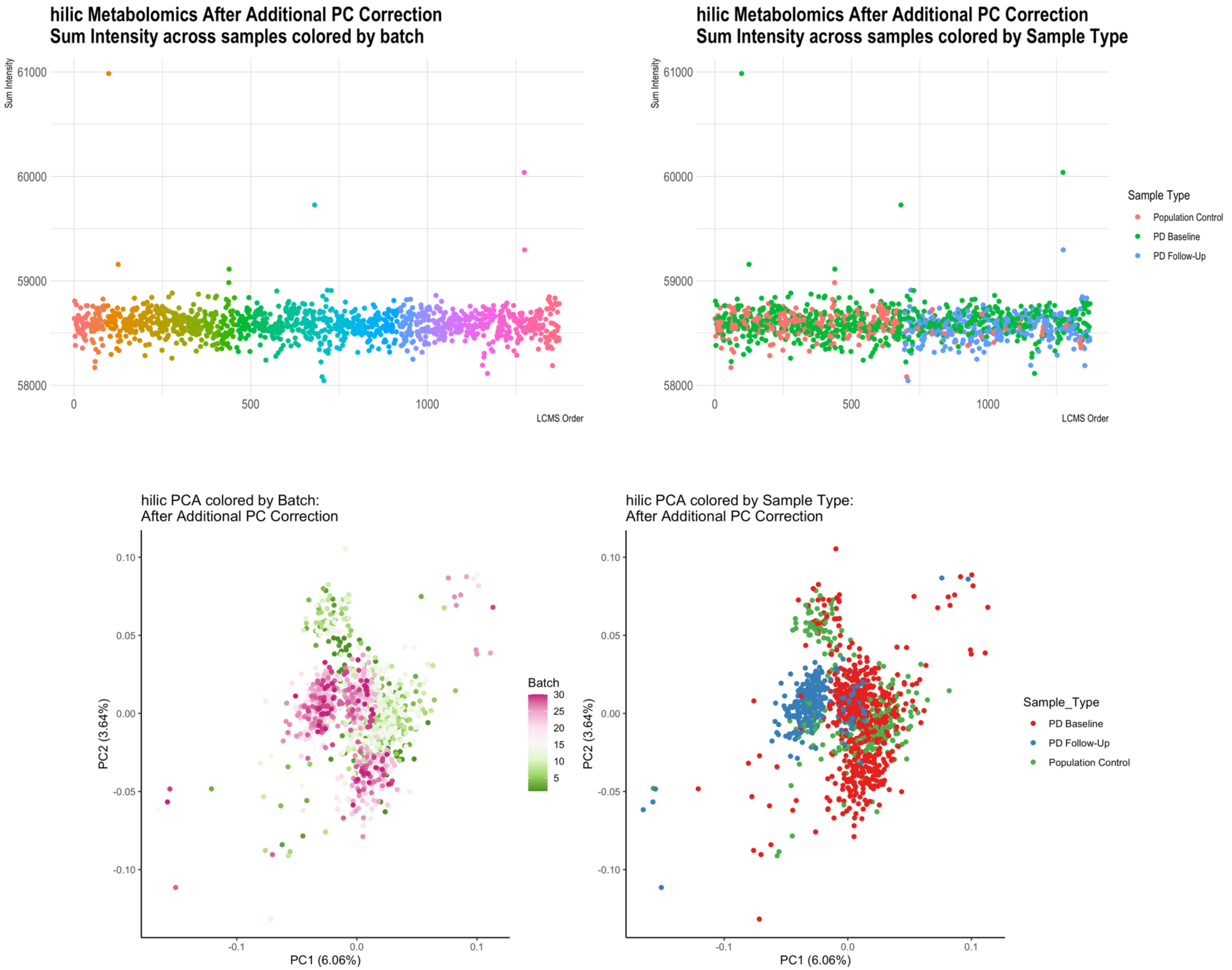


**Supplemental Figure 7. HILIC positive metabolomics data after processing:** Log transformation, quantile normalization, ComBat batch correction, and additional adjustment for unexplained PC.
